# Covid-19 related cognitive, structural and functional brain changes among Italian adolescents and young adults: a multimodal longitudinal case-control study

**DOI:** 10.1101/2023.07.19.23292909

**Authors:** Azzurra Invernizzi, Stefano Renzetti, Christoph van Thriel, Elza Rechtman, Alessandra Patrono, Claudia Ambrosi, Lorella Mascaro, Giuseppa Cagna, Roberto Gasparotti, Abraham Reichenberg, Cheuk Y. Tang, Roberto G. Lucchini, Robert O. Wright, Donatella Placidi, Megan K. Horton

## Abstract

Coronavirus disease 2019 (COVID-19) has been associated with brain functional, structural, and cognitive changes that persist months after infection. Most studies of the neurologic outcomes related to COVID-19 focus on severe infection and aging populations. Here, we investigated the neural activities underlying COVID-19 related outcomes in a case-control study of mildly infected youth enrolled in a longitudinal study in Lombardy, Italy, a global hotspot of COVID-19. All participants (13 cases, 27 controls, mean age 24 years) completed resting state functional (fMRI), structural MRI, cognitive assessments (CANTAB spatial working memory) at baseline (pre-COVID) and follow-up (post-COVID). Using graph theory eigenvector centrality (EC) and data-driven statistical methods, we examined differences in EC_delta_ (i.e., the difference in EC values pre- and post-COVID-19) and volumetric_delta_ (i.e., the difference in cortical volume of cortical and subcortical areas pre- and post-COVID) between COVID-19 cases and controls. We found that EC_delta_significantly between COVID-19 and healthy participants in five brain regions; right intracalcarine cortex, right lingual gyrus, left hippocampus, left amygdala, left frontal orbital cortex. The left hippocampus showed a significant decrease in volumetric_delta_between groups (p=0.041). The reduced EC_delta_ in the right amygdala associated with COVID-19 status mediated the association between COVID-19 and disrupted spatial working memory. Our results show persistent structural, functional and cognitive brain changes in key brain areas associated with olfaction and cognition. These results may guide treatment efforts to assess the longevity, reversibility and impact of the observed brain and cognitive changes following COVID-19.

## Introduction

SARS-CoV-2 infection claimed over 12.5 million lives worldwide resulting in the global COVID-19 pandemic with unique health, social and economic impacts^1^. Although primarily associated with respiratory symptoms, growing evidence suggests that COVID-19 is a multiorgan disease with impacts on the central nervous system, leading to various neurological outcomes such as headaches, anosmia, and altered cognition demonstrating the potential neurotoxic impact of the virus^2–4^. The underlying mechanisms through which COVID-19 impacts neural and cognitive functioning are not well established.

Magnetic resonance imaging (MRI) may prove useful in understanding, detecting, and monitoring neurobiological mechanisms underpinning COVID-19 related brain changes. Several MRI studies have shown significant anatomical brain changes between COVID-19 patients and healthy controls^5–7^. Changes include reduced gray matter thickness and decreased cerebral volume; specific brain areas have been reported in patients recovered from mild and severe forms of COVID-19. Reduced volume in the orbitofrontal cortex, parahippocampal gyrus, hippocampus and amygdala are associated with COVID-19 positivity^2, 5, 6^ and, in several cases, with cognitive deficits^5, 8^. Despite the many studies that investigate COVID-19 related structural brain changes, less is known about possible functional brain changes related to COVID-19. Resting-state fMRI (rs-fMRI) observes spontaneous (task-independent) signal fluctuations to investigate functional alterations in cortical and subcortical brain areas. Here, we leverage anatomical and rs-fMRI to better understand neural mechanisms underlying COVID-19 related brain changes. Our multimodal study includes MRI scans from COVID-19 positive cases and controls data collected before and after the infection.

While most studies of COVID-19 related brain and cognitive changes focus on severe infection or aged populations, most cases recorded worldwide were of mild to moderate illness in adolescents and young adults^8–11^. This population reported long-lasting symptoms including fatigue, headache, loss of concentration and memory problems^11^. Over 25% of mild COVID-19 cases reported visuoperceptual and visual organization deficits and linked these changes with structural brain alterations^8, 10^. In particular, spatial working memory (SWM), a critical cognitive function involving the ability to store and manipulate spatial information in the short term, was impacted by mild COVID-19 in adolescents, young adults and adults^8, 10^. Adolescence and young adulthood are key periods for brain growth and development^12–15^. Defining and strengthening of regional neurocircuitry and pathways between key brain areas such as amygdala, frontal lobe etc, is happening during adolescence. These brain areas play a crucial role in various cognitive and executive functions, including occipital, parietal and frontal lobes^12^. Further, adolescence and young adulthood are crucial periods for shaping behavior and interactions that rely on well-integrated cognitive mechanisms, executive and working memory functions^16–18^. These brain mechanisms are deeply shaped by social interactions that have been completely altered (i.e., social isolation and distancing) due to the COVID-19 pandemic. Thus, it is crucial to understand what are the implications of mild COVID-19 in adolescents and young adults which is the largest and understudied population of COVID-19 cases.

In this longitudinal multimodal MRI study, we compare pre- and post-anatomical, functional and cognitive outcomes in COVID-19 positive and healthy adolescents and young adults living in Lombardy, Italy, a global hotspot of COVID-19 during the pandemic.

## Materials and Methods

### Participants

The Public Health Impact of Metal Exposure (PHIME) study is a longitudinal cohort study of adolescents and young adults living in northern Italy. Details of the study have been described elsewhere^19, 20^. Inclusion criteria were: birth in the areas of interest; family residence in Brescia for at least two generations; residence in the study areas since birth. The exclusion criteria were: having a neurological, hepatic, metabolic, endocrine or psychiatric disorder; having clinically diagnosed motor deficits or cognitive impairment and having visual deficits that are not adequately corrected. Detailed description of this recruitment process and study design can be found in previous publications^20, 21^. Between 2016-2021, a convenience-based sample of 207 PHIME participants (53% female, ages 13-25 years) participated in a sub-study including multi-modal MRI scans and measures of memory and motor functions (Cambridge Neuropsychological Test Automated Battery (CANTAB))^22^. All participants satisfied the eligibility criteria for MRI scanning (i.e., no metal implants or shrapnel, claustrophobia, prior history of traumatic brain injury, and body mass index (BMI) ≤ 40). 202 participants completed the MRI and CANTAB tests. These data serve as the baseline in our study.

#### PHIME COVID-19 substudy

Beginning in March 2021, we invited all 207 participants to an in-person follow-up study (PHIME COVID-19) involving repeated identical MRI protocol and cognitive assessment as administered at baseline (PHIME study); 40 (19%) agreed to participate. Of these 40 participants, 16 reported having been infected with SARS-CoV-2 (COVID +) based on positive real-time reverse transcription polymerase chain reaction (RT-PCR) for SARS-CoV-2 RNA detection that was performed on a nasopharyngeal swab sample at the time of diagnosis. Based on literature suggesting SARS-CoV-2 antibodies titers decrease after 12 months^23, 24^, only participants with a positive RT-PCR test within 12 months of follow-up data acquisition were considered COVID + (Figure 1). Using these criteria, we identified 13 COVID+ and 27 COVID-subjects. Notably, four participants (1 COVID- and 3 COVID+ subjects that were positive but outside the 12 months threshold used to define COVID-19 status) reported mild COVID-19 symptoms and were treated as COVID- (see Table 1). Additional serological screening was performed only on COVID+ participants (available on 13 participants over 16 infected with SARS-CoV-2; Figure 1), retrospectively. Serum samples were tested using Euroimmun (Anti-SARS-CoV-2-NCP ELISA (IgG), Order No. El 2606-9601-2 G) assay with 80% sensitivity and 99.8%^25^. Results reported in Figure 1 are based on a ratio of specimen absorbance reported to the cut-off value defined by the manufacturer. None of the COVID-19 positive participants were hospitalized or suffered from pneumonia.

**Figure 1.**
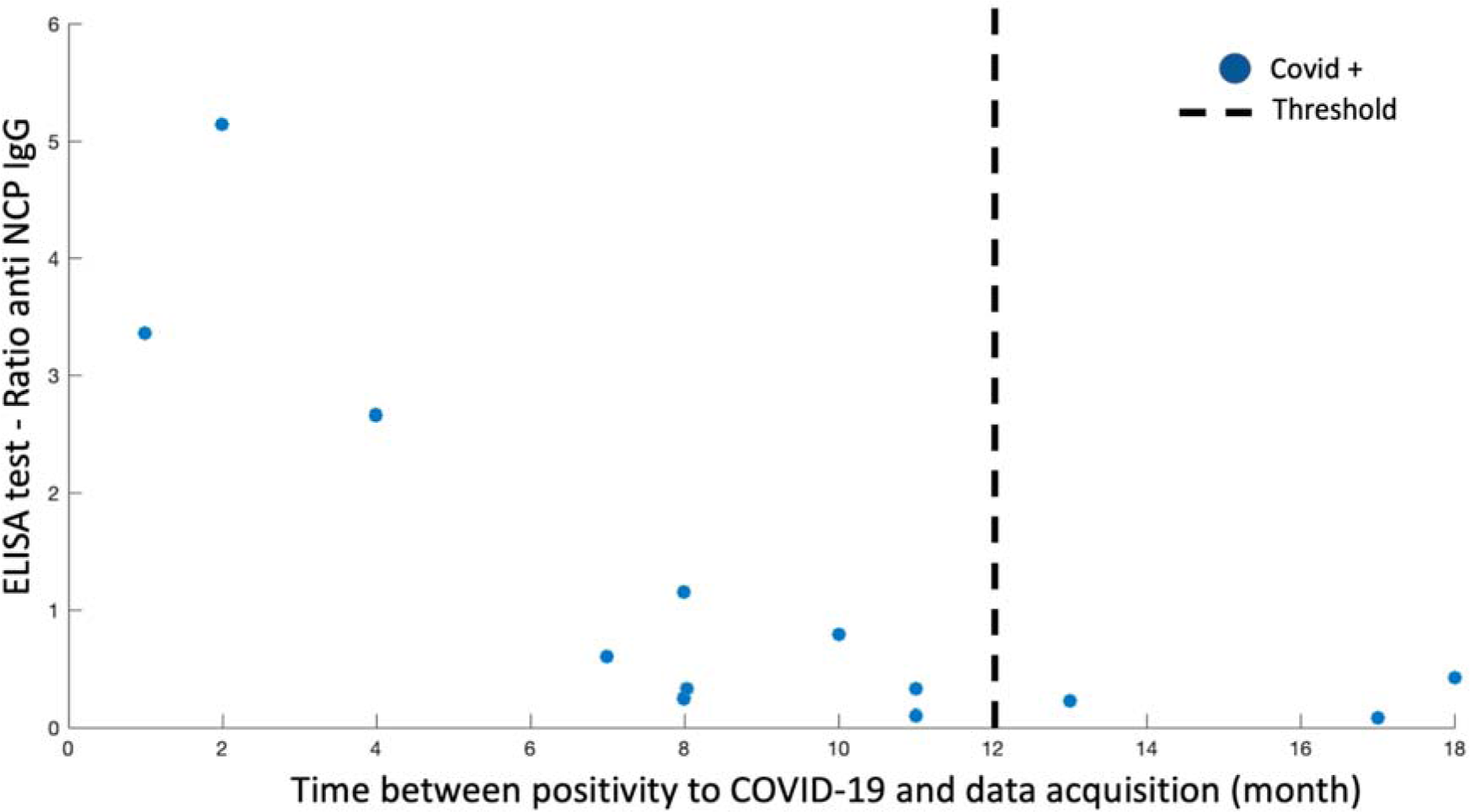
-COVID+ group definition. PHIME COVID-19 participants were included in the COVID+ group based on: 1) a positive RT-PCR for SARS-CoV-2 RNA detection and, 2) time of diagnosis within 12 months of data acquisition. The relation between the ratio of anti-SARS-CoV-2-NCP (IgG) detected using an ELISA assay and the time between a positive RT-PCR and data acquisition is reported in the figure for each single positive PHIME COVID-19 participant (blue dot). Three COVID+ participants are not included in the figure since the ELISA test is not available. Black dotted line represents the 12 month cut-off used as threshold for participant inclusion in the COVID+ group.

**Figure 2.**
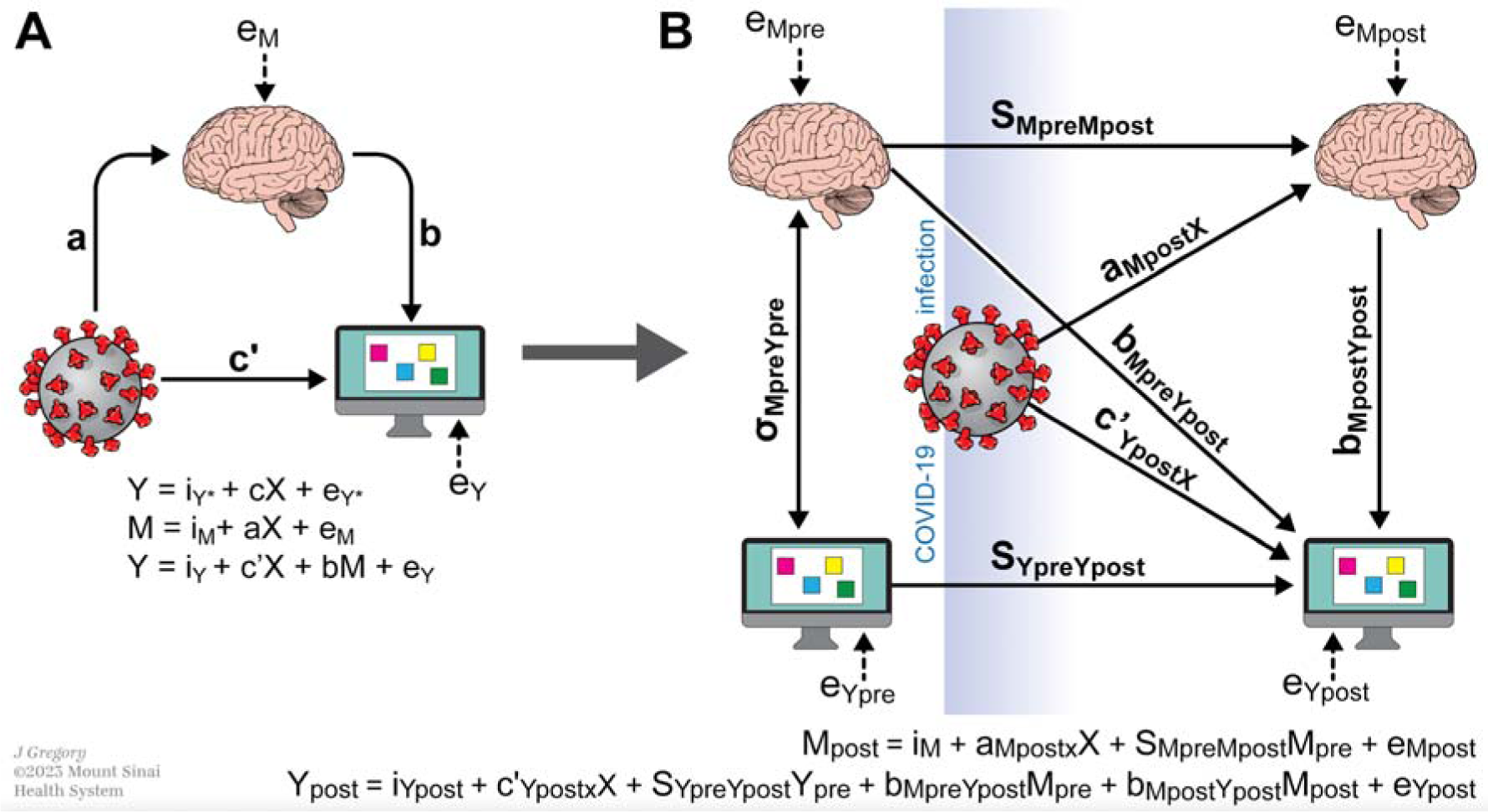
-Schematic depicting mediation models used. A. Classical mediation and regression models. The objective of the mediation analysis was to determine if the indirect effect (path a*b) was different from zero, suggesting that the mediating variable (i.e., brain metrics) altered the strength of the relationship between X and Y (i.e., COVID-19 status and spatial working memory metrics). Three linear regression equations were used to assess the mediation in the traditional cross sectional experimental design: Y regressed on X, M regressed on X, and Y regressed on both X and M . The letters a, b, c, and c’ refer to the regression coefficient estimates for each respective model, with e representing the error term. B. Given the longitudinal design of our study, we extended the traditional mediation analysis presented in panel A using a pre- and post-test control group design. Diagram includes: the pre-test covariance between mediator (M_pre_) and dependent variable (Y_pre_), σ_MpreYpre_; the effect of the mediator measured pre-COVID (M_pre_) on the mediator measured post-COVID (M_post_) (stability of mediator s_MpreMpost_); effect of the outcome measured pre-COVID (Y_pre_) on the outcome measured post-COVID (Y_post_) (stability of outcome s_YpreYpost_); the effect of M_pre_ on Y_post_ (cross-lagged relation, b_MpreYpost_); the effect of X on M_post_, a_MpostX_; the effect of X on Y_post_, c’_YpostX_ ;and the effect of M_post_ on Y_post_, b_MpostYpost_.

**Table 1.** -Sociodemographic and clinical characteristics of PHIME-COVID participants who were selected for the current study (N=40).

| Characteristics | COVID + | COVID - | p |
| --- | --- | --- | --- |
| **Number of subjects (n)** | 13 | 27 |  |
| **Sex (n, %)** |  |  | 0.291 |
| Male | 6 (46.15%) | 8 (30%) |  |
| Female | 7 (53.85%) | 19 (70%) |  |
| **Age at scan pre-COVID (years)** |  |  | 0.611 |
| (mean ± sd) | 20.44 ± 2.5 | 19.94 ± 2.3 | |
| **Age at scan post COVID (years)** |  |  | 0.721 |
| (mean ± sd) | 23.76 ± 2.82 | 24.1 ± 2.3 | |
| **Years between scans pre- and post-COVID (mean ± sd)** | 3.32 ± 1.44 | 4.12 ± 0.86 | 0.034 |
| **SWM pre-COVID**<br>(median, 1st and 3rd quartile) |  |  |  |
| "Between errors" | 13, [9, 23] | 10, [5.25, 17.75] | 0.369 |
| "Strategy" | 31.5, [29, 37] | 29, [27, 31] | 0.091 |
| **SWM post-COVID**<br>(median, 1st and 3rd quartile) |  |  |  |
| "Between errors" | 7, [0, 16.5] | 3, [0, 16] | 0.651 |
| "Strategy" | 31, [25.5, 33] | 28, [22, 30.75] | 0.131 |
| **COVID symptoms (n,%)** |  |  |  |
| Without symptoms | 4 (30.77%) | 23 (85.2%) | 0.001 |
| With symptoms | 9 (69.23%) | 4 (14.8%)* |  |
| **COVID vaccine (n,%)** |  |  |  |
| Without vaccine | 2 (15.4%) | 0 | 0.001 |
| With vaccine | 11 (84.6%) | 27 (100%) |  |

**Note:** Mean, standard deviation (sd), range (minimum and maximum values), and percentage (%) are reported. P-values were derived using Student's *t*-tests and Wilcoxon rank sum test for continuous variables (if the variable was normally distributed or not, respectively) and tests for categorical variables to assess the differences between COVID+ and COVID-.
\*COVID-PHIME participants reported mild-COVID symptoms. These participants reported symptoms outside of the 12 months threshold used to define COVID positivity.

Written informed consent was obtained from parents and young adults, participants < 18 years provided written assent. Study procedures were approved by the Institutional Review Board of the University of California, Santa Cruz and the ethical committees of the University of Brescia, and the Icahn School of Medicine at Mount Sinai.

### Neuropsychological assessment

Trained neuropsychologists administered the standard Cambridge Neuropsychological Test Automated Battery (CANTAB)^22^ to assess domains of cognitive, memory and motor functioning. To test our hypothesis on the impact of COVID-19 on the brain and working memory functions, we used the CANTAB Spatial Working Memory (SWM) test. This test provides a comprehensive evaluation of an individual’s ability to retain and manipulate spatial information in working memory. The SWM task detects frontal lobe activity and executive dysfunctions. The participants are shown different colored squares (boxes) on a screen and asked to identify yellow tokens within a box. Participants are instructed to search for tokens by opening boxes (touching each box on the screen) and advised to not return to the same box that already contained a token. These tokens are then used to automatically fill an empty column located on the right side of the screen. The complexity of the task gradually increases as the number of boxes to be searched reaches a maximum of twelve. To discourage the use of stereotypical search strategies, the color and position of the boxes are changed between trials. Several outcome measures are provided by SWM test, including error rates, instances of selecting empty boxes, revisiting boxes that already contain a token, as well as measures of strategy implementation and latency. Here, we focused on the “between error”, number of times that a participant selected a box that already contained a token, and “strategy”, number of times that a participant selected a new and different box looking for a new token. Both variables are available for 39 participants who completed the CANTAB.

### MRI and fMRI data acquisition

Magnetic resonance imaging (MRI) and functional MRI (fMRI) data acquisition was performed on a high-resolution 3-Tesla SIEMENS Skyra scanner using a 64-channel phased array head and neck coil, at the Neuroimaging Division of ASST Spedali Civili Hospital of Brescia. For each participant, a high-resolution 3D T1-weighted structural scan was acquired using a MPRAGE sequence (TR =2400 ms, TE= 2.06 ms, TI=230 ms, acquisition matrix=256×256 and 224 sagittal slices with final voxel size=0.9 mm^3^). Fifty contiguous oblique-axial sections were used to cover the whole brain where the first four images were discarded to allow the magnetization to reach equilibrium. For each subject, a single 10-minute continuous functional sequence using a T2*weighted echo-planar imaging (EPI) sequence (TR=1000 ms, TE=27 ms, 70 axial slices, 2.1 mm thickness, matrix size 108×108, covering the brain from vertex to cerebellum) was acquired. During resting-state scans, lights of the MRI room were off and participants were instructed to stay awake, relax and daydream (not think about anything) with their eyes open. They were presented with an image of a night skyline figure projected on a MRI compatible monitor. Padding was used for comfort and reduction of head motion. Earplugs were used to reduce noise. Data were read by a board-certified radiologist to determine the quality and possible incidental findings - no findings were reported.

#### MRI and fMRI data analyses

Image pre-processing, eigenvector calculations, and statistical analyses were performed using FreeSurfer software (v7.1.1; https://surfer.nmr.mgh.harvard.edu/), SPM12 (Wellcome Department of Imaging Neuroscience, London, UK), Brain Connectivity toolbox ^26, 27^ and customized scripts, implemented in MatLab 2016b (The Mathworks Inc., Natick, Massachusetts) and R (v3.4).

#### MRI processing and analysis

T1-weighted scans were preprocessed using FreeSurfer to perform cortical and subcortical reconstruction and segmentation^28–31^. The standard cross-sectional pipeline available in Freesurfer v. 7.1.1 including intensity normalization, automated topology corrections, and automatic segmentation of cortical and subcortical brain areas was applied to each subject. Total intracranial volume (TIV) was also extracted^31^.

#### fMRI Image preprocessing

For each subject, the structural MRI was co-registered and normalized against the Montreal Neurological Institute (MNI) template and segmented to obtain white matter (WM), gray matter (GM) and cerebrospinal fluid (CSF) probability maps in the MNI space. FMRI data were spatially realigned, co-registered to the MNI-152 EPI template and subsequently normalized utilizing the segmentation option for EPI images in SPM12. All normalized data were denoised using ICA-AROMA^32^. Additionally, spatial smoothing was applied (8 millimeters) to the fMRI data. As a further quality check of fMRI data, large head motion in any direction or rotation (> 3mm or 3°) was used as exclusion criteria in our study - no participants were excluded in this study. No global signal regression was applied.

Based on the Harvard-Oxford^33^ atlas, 111 regions of interest (ROI; 48 left and 48 right cortical areas; 7 left and 7 right subcortical regions and 1 brainstem) were defined. In the Harvard-Oxford atlas, brain areas were defined using T1-weighted images of 21 healthy male and 16 healthy female subjects (ages 18-50). The T1-weighted images were segmented and affine-registered to MNI152 space using FLIRT (FSL), and the transforms were then applied to the individual brain areas’ labels. Finally, these were combined across subjects to form population probability maps for each ROI^33^. For each ROI, a time-series was extracted by averaging across voxels per time point. To facilitate statistical inference, data were “pre-whitened” by removing the estimated autocorrelation structure in a two-step generalized linear model (GLM) procedure^34, 35^. In the first step, the raw data were filtered against 6 motion parameters (3 translations and 3 rotations). Using the resulting residuals, the autocorrelation structures present in the data were estimated using an Auto-Regressive model of order 1 (AR(1)) and then removed from the raw data. Next, the realignment parameters, white matter (WM) and cerebrospinal fluid (CSF) signals were removed as confounders on the whitened data.

#### Local and global functional connectivity analysis

<u>Eigenvector centrality mapping (ECM)</u> is a measure to spatially characterize connectivity in functional brain imaging by attributing network properties to voxels^26, 36–38^. The ECM method builds on the concept of eigenvector centrality, which characterizes functional networks active over time and attributes a voxel-wise centrality value to each ROI. Such a value is strictly dependent on the sum of centrality properties of the direct neighbor ROI within a functional network. In our study, fast ECM (fECM)^39^ toolbox was used to estimate voxel-wise eigenvector centralities (EC) from the time course data extracted based on the Harvard-Oxford ROIs definition per subject. ECM is estimated from the adjacency matrix, which contains the pairwise correlation between the ROIs. To obtain a real-valued EC value, we added +1 to the values in the adjacency matrix. Several EC values can be attributed to an individual node by the ECM method^39^, but only the eigenvector with the highest eigenvalue (EV) will be used for further analyzes for each node. The highest EV values were averaged across subjects at group level.

<u>Functional connectivity (FC)</u> was computed for each participant applying the pairwise temporal Pearson correlation between ROIs and a Fisher’s z-transformation. The ROI’s z-values were averaged across participants. The difference in median FC-values pre- and post-COVID-19 (FC_delta_) is calculated and used to compare FC-values between groups (COVID+ vs COVID-).

### Statistical analyses

#### Descriptive statistics

Pairwise Student t-tests with Welch’s correction for continuous variables were used to examine differences in clinical and demographic characteristics across the groups.

Kolmogorov-Smirnov test was used to check the normality of the brain structural data and then, a linear regression was applied to adjust for sex, years between pre- and post-COVID scans and pre-COVID age. Pairwise Student t-tests were used to examine differences in brain structural metrics and TIV pre- and post-COVID-19 between groups.

#### Permutation statistics

We quantify possible functional hubs by comparing the EC_delta_ (i.e., the difference in EC values pre- and post-COVID-19) values across groups using a family-wise error corrected (FWE) permutation test. Permuted labels based on group definitions (COVID + and COVID -) were repeated 1000 times per subject. Only ROIs with EC values that differ significantly were considered functional hubs. For both local and global functional connectivity analysis (EC and FC metrics), the FWE correction was applied for the number of group level comparisons and for the total number of ROIs analyzed. Linear regressions were applied to adjust for sex, years between pre- and post-COVID scans, and pre-COVID age. Only p-values adjusted and FWE corrected are reported.

#### Mediation analysis

To test the mediating role of brain metrics between COVID-19 and spatial working memory accounting for the longitudinal design of the study, a two-waves mediation analysis was applied (Figure 1, panel B)^40, 41^. This extension of the traditional mediation analysis (Figure 1, panel A) accounts for longitudinal mediated effects of data^40^. A linear regression model was applied to test the effect of COVID-19 on the logit-transformed mediator (each brain area) measured at the second visit and adjusted for the baseline measurement. A negative binomial and a linear regression were considered for the full models where the between errors and the strategy scores (representing the SWM metrics) were considered as outcomes, respectively. Only brain areas identified as hubs in the permutation analysis and with significant association with COVID-19 were included in this analysis. Models were adjusted for sex, years between pre- and post-COVID scans and pre-COVID age. The statistical significance level was set to 5% and all tests were two-sided. Statistical analyses were performed using R (Version 4.3.1), the mediation analysis was conducted using the mediation^42^ R package (Version 4.5.0).

## Results

### Demographic characteristics

Table 1 reports the clinical and demographic characteristics of the 40 COVID-PHIME participants included in this study stratified by COVID-19 status; positive (COVID +) and negative (COVID -). Participants were adolescents or young adults at the time of the first imaging data acquisition (20.44 ± 2.5 years) and the majority were females (65%). Years between pre- and post-COVID assessments, COVID symptoms and vaccine status significantly differ between groups. A complete overview of COVID symptoms reported by COVID+ participants can be found in Supplementary Materials (Table S1).

### Structural and functional differences between COVID+ and COVID-

EC values in permutation tests (COVID+ vs. COVID-) revealed five functional hubs where EC_delta_ values differed significantly between COVID+ and COVID-groups including: right intracalcarine cortex, right lingual gyrus, left hippocampus, left amygdala, and left frontal orbital cortex (Table 2, Figure 3A and 3B).

**Table 2.** -Statistical differences in EC_delta_ values between COVID+ and COVID-participants.

| Covid + versus Covid - |  |  |
| --- | --- | --- |
| Brain region (ROIs) | Hemisphere | p* |
| Intracalcarine Cortex (CAL) | R | 0.0444 |
| Lingual Gyrus (LING) | R | 0.0405 |
| Hippocampus (HIP) | L | 0.0439 |
| Amygdala (AMYG) | L | 0.0408 |
| Frontal Orbital Cortex (FOC) | L | 0.0286 |

Table 2 reports the full name of the brain regions interest (ROIs), ROI's hemisphere, ROI's abbreviation and p-values of the identified ROIs for which $EC_{\text{delta}}$ differed significantly between PHIME participants with and without COVID-19 (i.e., functional hubs). Only p-values FWE corrected, adjusted for sex, years between scans and age at the first visit (pre-COVID) are reported.

**Figure 3.**
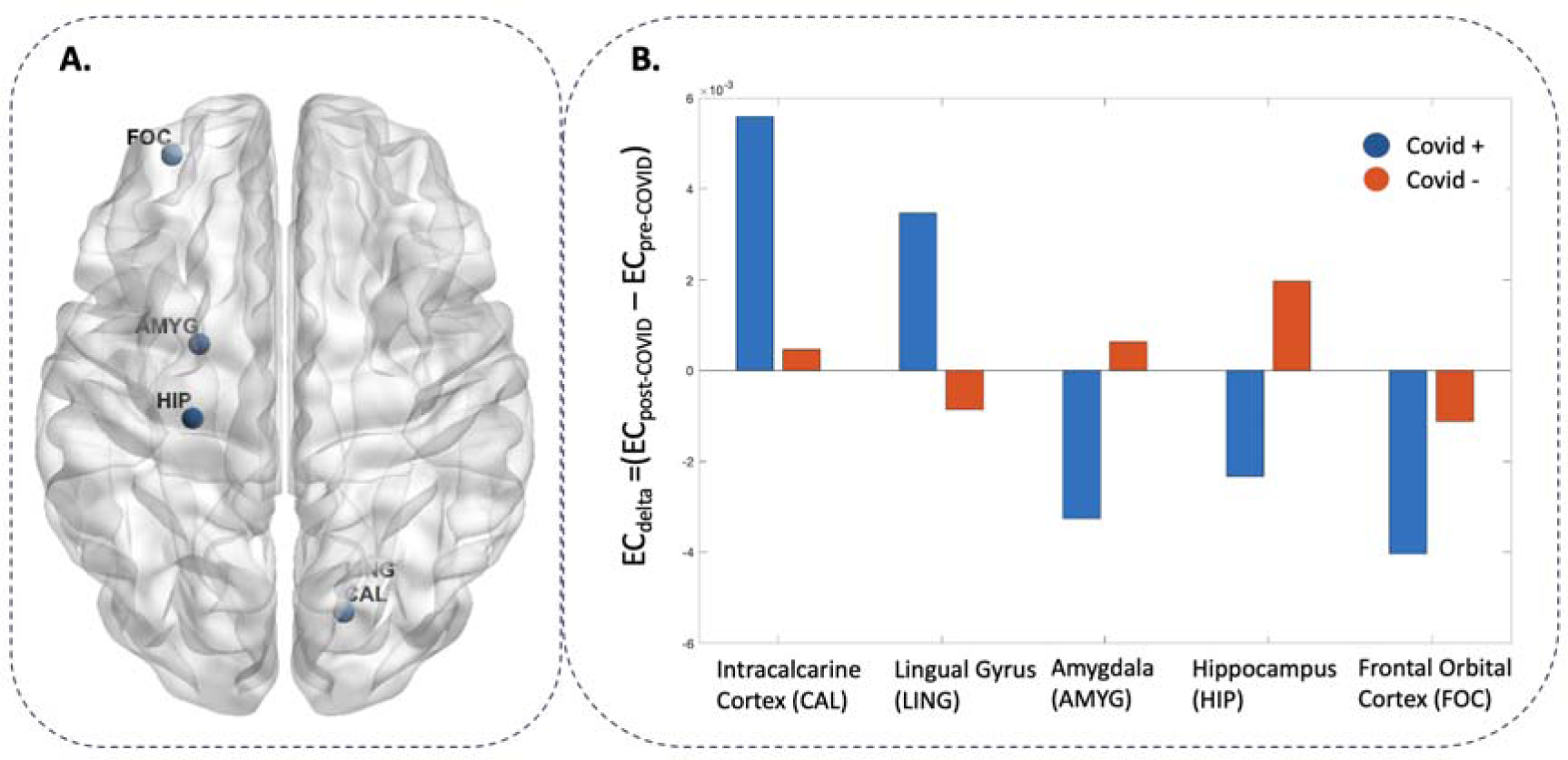
-Functional hubs differ in EC_delta_ between COVID+ and COVID-participants. Panel A reports the identified ROI for which EC_delta_ (differences in eigenvector centrality (EC) between pre- and post-COVID) differed significantly between PHIME participants with and without COVID-19 (i.e., functional hubs). Panel B reports the average EC_delta_ values for each functional hub identified. Blue bars indicate EC values of COVID+ participants, while orange bars indicate COVID-participants.

For brain areas identified as functional hubs, structural differences between groups were investigated using volumetric values adjusted for TIV (Table 3, Figure 4). Only the left hippocampal volume was reduced in COVID+ compared to healthy participants (p=0.034, Table 4).

**Table 3.** -Statistical differences in Volumetric_delta_ between COVID+ and COVID-participants.

| **Covid + versus Covid -** |  |  |
| --- | --- | --- |
| **Brain region (ROIs)** | **Hemisphere** | **p** |
| Intracranial volume | - | 0.311 |
| Intracalcarine Gyrus | R | 0.296 |
| Lingual Gyrus | R | 0.212 |
| Hippocampus | L | 0.034 |
| Amygdala | L | 0.404 |
| Frontal Orbital Cortex | L | 0.859 |

Table 3 reports the statistical differences in Volumetric<sub>delta</sub> (difference in volumetric metrics between pre- and post-COVID) between PHIME participants with and without COVID-19. Only brain areas that are previously identified as functional hubs (i.e., brain areas for which EC<sub>delta</sub> differed significantly between PHIME participants with and without COVID-19) are included in this analysis. The table reports the full name of the brain regions interest (ROIs), ROI's hemisphere and p-values. Only p-values adjusted for sex, years between scans, age at the first visit (pre-COVID) and total intracranial volume are reported.

**Table 4.** – Two-waves mediation analysis of the effect of COVID-19 on the cognitive scores (between errors and strategy metric obtained from SWM test) considering the brain area selected as functional hub (EC values) as mediators.

| **Direct effects (COVID-19 -> SWM)** |  |  |  |  |
| --- | --- | --- | --- | --- |
| *Brain Area* | *SWM metrics* | *Estimate* | *Confidence Interval* | *P-values* |
| Amygdala, left | Strategy | 0.8 | [-2.7; 4.1] | 0.656 |
| Amygdala, left | Between errors | -21.6 | [-89.4; 4.7] | 0.134 |
| **Mediated or Indirect effects (COVID-19 -> Brain -> SWM)** |  |  |  |  |
| *Brain Area* | *SWM metrics* | *Estimate* | *Confidence Interval* | *P-values* |
| Amygdala, left | Strategy | 0.3 | [-1.1; 1.8] | 0.602 |
| Amygdala, left | Between errors | 18.7 | [0.8; 82.6] | 0.028 |
| **Total effects (direct + mediated effects)** |  |  |  |  |
| *Brain Area* | *SWM metrics* | *Estimate* | *Confidence Interval* | *P-values* |
| Amygdala, left | Strategy | 1.2 | [-2.2; 4.2] | 0.506 |
| Amygdala, left | Between errors | -2.9 | [-35.0; 27.5] | 0.752 |

Two-waves mediation analysis performed to assess the direct, indirect and total effects of COVID-19 on SWM. EC brain values at baseline (pre-COVID), sex, years between pre- and post-COVID scans, pre-COVID age and SWM at baseline were used as covariates in the models. Only brain areas identified as hubs in permutation analyzes were entered as outcomes in this analysis. Then mediation analyses were performed using a pre-post test design. The moderating effects of brain metrics (eigenvector centrality value of a single brain area defined using the Harvard-Oxford atlas between COVID-19 and cognitive domains, in this case SWM).

**Figure 4.**
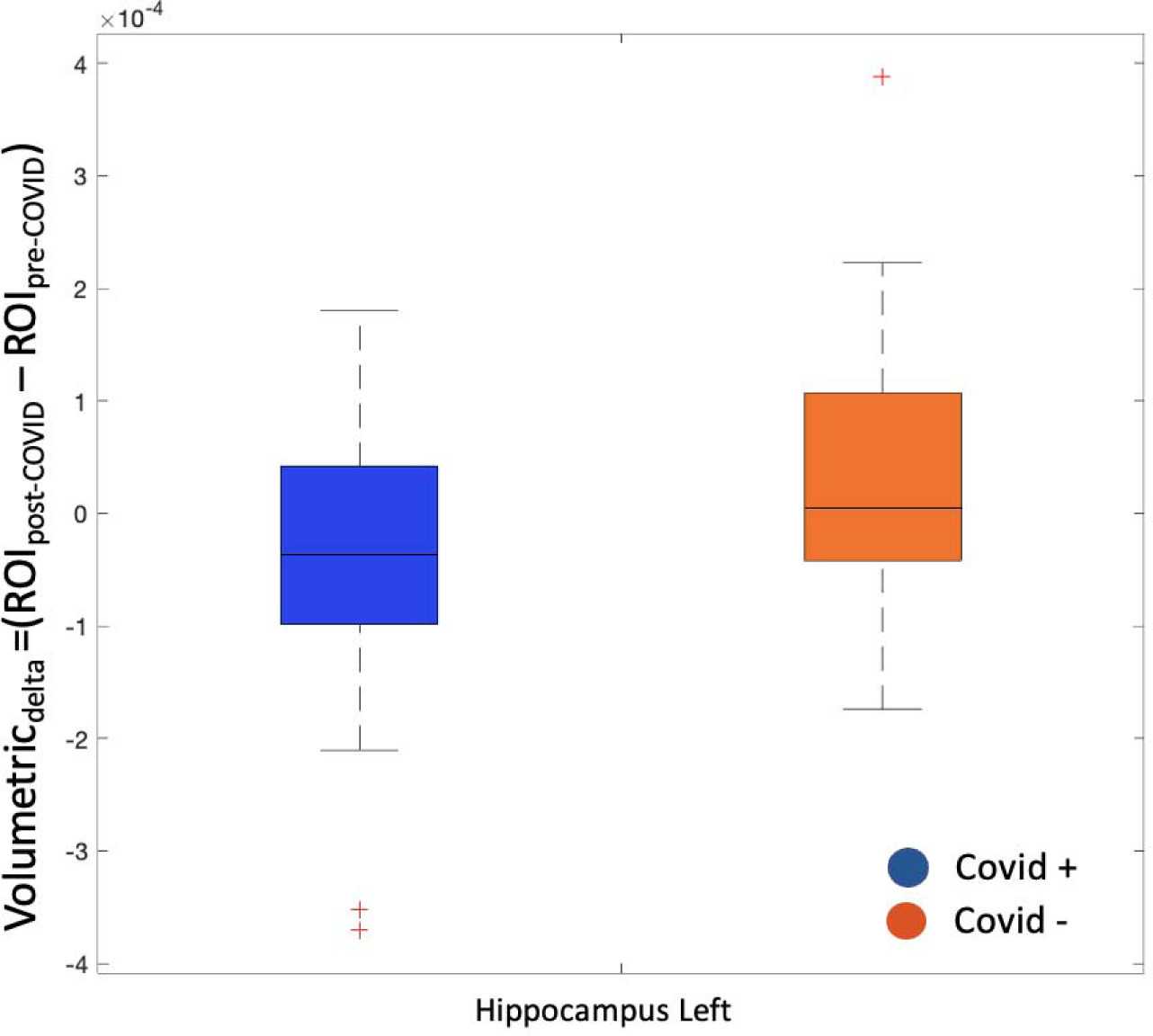
-Volumetric difference in the hippocampus between COVID+ and COVID-participants. The left hippocampal Volumetric_delta_ (difference in volumetric metrics between pre- and post-COVID) statistically differs between PHIME participants with and without COVID-19. The blue box indicates volumetric values of COVID+ participants, while the orange box indicates volumetric values of COVID-participants. Black lines indicate the median value.

We also compared whole brain connectivity between COVID+ and healthy participants by plotting the averaged FC scores across all ROIs pre- and post-COVID and the difference in functional connectivity metric between pre- and post-COVID (Figure S1, supplementary material). No significant differences were found (FC_delta_ p= 0.786).

### Mediation analysis

We observed significant associations between COVID-19 status and brain metrics in only one of the five functional hubs; the left amygdala (p-values for interaction= 0.032). Mediation analyses were performed to quantify possible indirect effects of the brain on the association between COVID-19 and spatial working memory in the functional hubs significantly associated with COVID-19. Results indicate that the left amygdala mediated the association between COVID-19 and SWM “between errors” (p-values for indirect effect= 0.028). No significant associations were found for SWM “strategy” SWM.

## Discussion

Leveraging an ongoing longitudinal study of adolescents and young adults in Lombardy, we used MRI, rs-fMRI and cognitive data to investigate local connectivity and structural differences associated with mild COVID-19. We identified connectivity differences in five cortical and subcortical brain areas between participants with and without mild SARS-CoV-2 infection. Additionally, we observed reduced volume in the left hippocampus of COVID+ compared to COVID-subjects. We observed a significant association between mild SARS-CoV-2 infection and functional metrics in one subcortical brain area, the left amygdala. Previous literature linked COVID-19 to structural and functional brain changes and cognitive deficits^6, 43–45^. Our results suggest that decreased functional connectivity in the amygdala competitively mediated the COVID-19 associated disruption in cognitive function, specifically spatial working memory. These results add to the previous literature documenting structural and cognitive changes associated with COVID-19 and further contribute to our understanding of the neurobiological underpinnings of mild COVID-19 in adolescents and young adults.

One out of four mild COVID-19 subjects reported persistent deficits in higher cognitive functions like concentration, memory, visuospatial and visuoconstructive abilities^10, 46^. Our study focused on the spatial working memory (SWM), a critical cognitive function that involves the capacity to store and manipulate spatial information in the short term. Studies suggest SWM is impacted by COVID-19^8, 10^ and linked with brain changes^8^. Our results suggest functional connectivity in one key area for cognitive functions, the amygdala, competitively mediates the association between COVID-19 positivity and SWM (Table 4), as only the indirect effect is statistically significant^47–49^. The amygdala is involved in the integration, interpretation, and storage of the visual information perceived, as well as in short memory and executive functions^50^. Our results are consistent with previous studies reporting deficits in several cognitive functions, including visual and verbal domain of short term memory, attention and memory associated with COVID-19^8, 51–53^. Future studies are needed to comprehensively understand the impact of COVID-19 on cognitive functions that are key for everyday life activities (i.e., movement planning, driving, etc.) and their link with the underlying neural mechanisms.

No significant overall functional connectivity disruption nor brain volume reduction were observed among participants who tested positive for SARS-CoV-2 infections (Figure S1). These results align with previous studies in non-hospitalized elderly populations^5, 8, 46, 54, 55^ suggesting that mild COVID-19 may impact specific brain cortical and subcortical areas rather than the whole-brain, regardless of the age at diagnosis.

Combining rs-fMRI data together with a graph-theory based method, we identified five functional hubs - the right intracalcarine cortex, the right lingual gyrus, the left frontal orbital cortex, the left hippocampus and left amygdala. Notably, three of these functional hubs, frontal orbital cortex, hippocampus and amygdala, undergo significant growth, maturation and connectivity changes across adolescence and young adulthood^56^. Located in the primary visual cortex, the intracalcarine cortex and lingual gyrus process, integrate and interpret the visual stimuli^57, 58^. Connected with subcortical areas such as the hippocampus and amygdala, these areas are involved in the perceptual learning and recognition of complex, non-conscious visuospatial sequences, visual memory and memory performance^59–61^. Altered connectivity between these cortical areas and the hippocampus have previously been associated with poor recollection, memory performance^62, 63^ and cognitive decline^64–66^. We observed increased functional connectivity in the intracalcarine cortex and lingual gyrus together with unbalanced connectivity coupling with subcortical areas combined with poor SWM functions only in the positive COVID-19 group (Figure 3b, Table 2). Our findings are in line with previous studies reported in structural and functional alterations in lingual gyrus and intracalcarine cortex throughout different phases of SARS-CoV-2 infection^8, 67–70^ and further support the role of these areas in low-level perceptual learning and memory performance.

Beside containing the secondary taste and olfactory areas, the orbitofrontal cortex plays a key role receiving and integrating sensory information (i.e., smell, taste and visual) and in different cognitive functions (i.e., attention, emotional and social behavior, decision making and conflict-error monitoring)^71–73^, which have been highly impaired in COVID-19 subjects. Highly connected with temporal lobe areas (i.e. amygdala, hippocampus and entorhinal cortex) and with other cortical regions like the cingulate cortex, the orbitofrontal cortex has been indicated as one of the main brain areas involved in the neuroinvasive pathway of SARS-CoV-2 infection^74–77^. Decreased functional connectivity observed in this areas only in the positive COVID-19 group (Figure 3b, Table 2) further support this theory together with previous studies that identified structural, functional, and metabolic changes in the orbitofrontal regions in individuals with acute, mild and long-term COVID-19, spanning different age groups and including both hospitalized and non-hospitalized participants^5, 78–83^. Future studies should further investigate this neuroinvasive pathway of SARS-CoV-2 by including additional olfactory structures (i.e., olfactory bulbs) and detailed symptoms report.

Two out of the five identified functional hubs were located in the subcortical brain structures (Figure 3). Closely connected with olfactory bulbs and orbitofrontal areas, the left amygdala is the only brain area showing a significant association with COVID-19 positivity and hypo-connectivity among the COVID+ group defined as decreased functional connectivity (Figure 3b, Table 2). Further, we found structural and functional changes in the left hippocampus positively associated with COVID-19 status (Figure 3, 4; Table 2,3). Interestingly, only this subcortical brain area shows a significantly reduced cortical volume in our COVID+ group. Linked with cognitive, spatial and working memory functions as well as learning, hippocampal areas have been severely impacted by COVID-19 leading to alterations in its cortical volume, microstructure and functional connectivity^56, 84–87^ across different age groups and COVID-19 phases.

Our findings are in line with previous studies that have reported structural and functional alterations in amygdala and hippocampus throughout different phases of SARS-CoV-2 infection and among populations^5, 45, 88–90^ that faced unique social stressors (i.e., social isolation, lockdown), during COVID-19 pandemic. However, it is still unclear if these brain changes are due to SARS-CoV-2 infection or to the psychosocial stressors experienced during COVID-19 pandemic triggering resilience effects^10^. Both amygdala and hippocampal areas are key in predicting vulnerability and resilience to stress-related disorders like post-traumatic stress disorder (PTSD)^91–93^. These areas are also heavily involved in personality, emotional, and behavioral regulation^94, 95^, fear processing and fear conditioning^96, 97^, and the formation of memories related to stressful events^98^. Future studies should include stress metrics to understand the involvement of amygdala and hippocampus in psychosocial stress responses, shedding light on their potential as neural basis for individual variations in stress reactivity, vulnerability^89^ and predisposition to stress-related disorders in adolescents and young adults.

Our work identified specific cortical and subcortical brain areas affected by mild COVID-19 in adolescent and young adults, achieved through the use of rs-fMRI, MRI, and cognitive data using a reliable functional-connectivity based approach. A notable strength of our research is the availability of baseline imaging and cognitive tests conducted before and after infection. By leveraging the longitudinal nature of our multi-modal study, which includes high quality pre- and post-morbidity imaging and cognitive assessments, we were able to assess the impact of the COVID-19 pandemic in the adolescent and young adult population. Moreover, we employed graph-based network metrics derived from rs-fMRI to investigate alterations in both brain and cognitive functions resulting from mild COVID-19. These metrics offer the advantage of being unconstrained by prior seed selection of hypothesis-driven approaches, thus proving a comprehensive and unbiased assessment of the observed brain changes.

Limitations of our study design include the small sample size and lack of an external control group outside the PHIME cohort. Our small sample size prohibited seeing total effects of COVID-19 on SWM metric that might be attenuated by the negative no-significant direct effect of COVID-19 on SWM metric estimated. A larger sample size might improve statistical power and allow us to identify additional functional and structural brain areas associated with COVID-19 as well as a direct effect of COVID-19 on cognitive metrics. In addition, our cohort lacks diversity and consisted only of white participants aged between 13-25 years, meaning that the findings cannot be generalized to other populations. While it would be beneficial to repeat our analysis in a larger dataset, there is no such dataset that includes multi-modal imaging, cognitive and COVID-19 symptomatology collected in Italy, specifically, in the global hotspot of COVID-19. In addition, the average time interval between MRI scans is long (^~^3 years) in comparison to the average time between documented infections and the second MRI scan (12 months at maximum). The relatively long time between MRI studies increases the potential for confounding factors, such as age-related changes in functional or cognitive functions. An additional longer-term follow-up study will assess the reversibility of the observed post-infectious changes and disentangle them from the above mentioned confounding variables. On the basis of the study frame, SARS-CoV-2 infections among participants were presumably caused by one of the many SARS-CoV-2 variants (alpha, beta or gamma). Further analysis of these data, including correlation with olfactory symptoms could provide more insight into the vulnerability of subgroups.

To conclude, this is a multimodal longitudinal case-control study using cognitive assessment, rs-fMRI and MRI data to provide novel insights into the underlying neural and cognitive mechanisms of adolescents and young adults living in a global hotspot of COVID-19 during the pandemic. Our results show persistent functional and structural changes in specific brain areas in participants COVID-19 positive. These changes involved gray matter volume and FC connectivity in cortical and subcortical areas previously shown to be associated with mild and severe COVID-19. These changes are associated with COVID-19 status, as well as with a drop in cognitive functions, in particular SWM metrics. Future studies to assess the longevity and reversibility of the observed brain and cognitive changes following infections are still needed to advance our understanding in future development of post-COVID infection and to help intervention and treatment.

## Data Availability

All data produced in the present study are available upon reasonable request to the author

## Acknowledgements

Illustration by Jill K. Gregory. Used with permission of ©Mount Sinai Health System. The authors would like to acknowledge Carsten Watzl and Doris Urlaub for the performing the ELISA analysis and support from the National Institutes of Environmental Health Sciences (NIEHS) grants numbers R01 ES019222, R01 ES013744, P30ES023515, and the European Union through its Sixth Framework Programme for RTD (contract number FOOD-CT-2006-016253).

Conflict of Interest: None.

## Supplementary Materials

**Figure S1.**
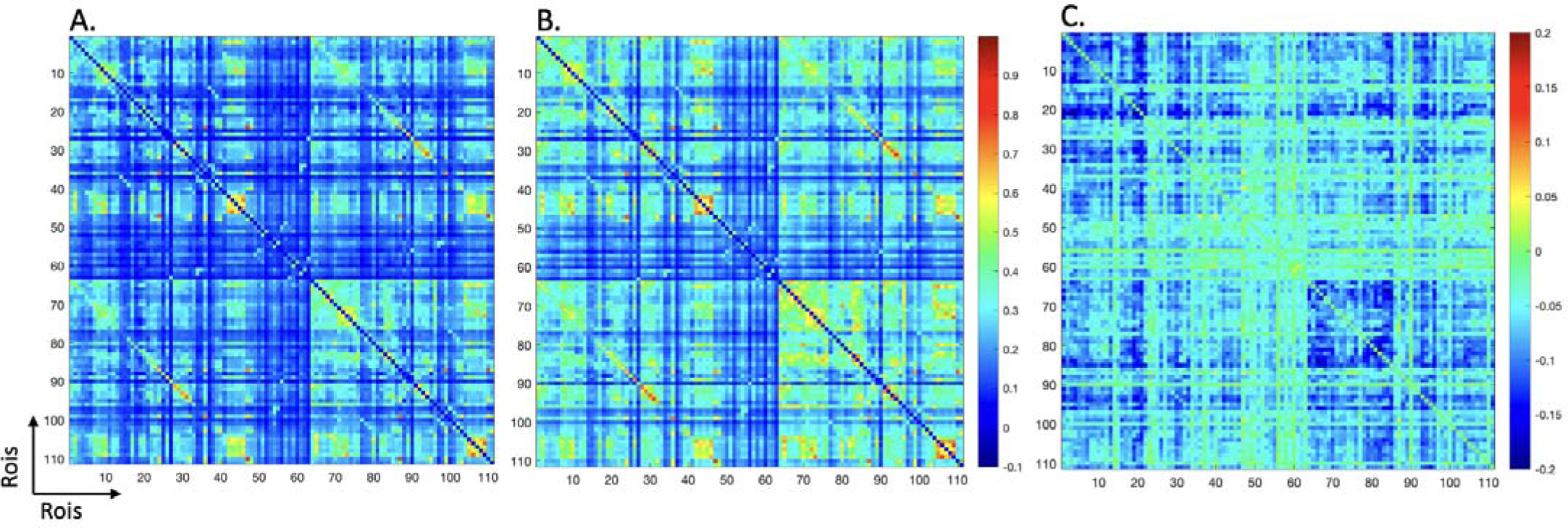
-Whole brain functional connectivity matrices, based on the Harvard-Oxford atlas. For each timepoint, first all individual functional connectivity (FC) matrices were calculated by applying Fisher’s r-to-z transformation, which were then averaged across participants. Panels a, b, c and d show the mean inter-ROI correlation matrices for pre-COVID (baseline), post-COVID and estimated FC_delta_(i.e., the difference in FC values pre- and post-COVID-19), respectively.

**Table S1.** -COVID symptoms reported by COVID+ participants.

| **COVID symptoms (n,%)** | **Symptomatic** | **Asymptomatic** |
| --- | --- | --- |
| *Fever* | 6 (46.15 %) | 7 (53.85 %) |
| *Respiratory infection (upper tract)* | 2 (15.38 %) | 11 (84.62 %) |
| *Respiratory infection (lower tract)* | 0 | 13 (100 %) |
| *Anosmia (loss of sense of smell)* | 3 (23.1 %) | 10 (76.92 %) |
| *Ageusia (loss of sense of taste)* | 3 (23.1 %) | 10 (76.92 %) |
| *Headache* | 3 (23.1 %) | 10 (76.92 %) |
| *Tiredness* | 5 (38.46 %) | 8 (61.54 %) |
| *Muscle aches* | 4 (30.77 %) | 9 (69.23 %) |
| *Gastrointestinal disorders* | 0 | 13 (100 %) |

**Note:** Number of subjects and percentage (%) are reported for COVID+ participants.

